# Assessment of knowledge retention of healthcare staff through telephonic interview after training for rotavirus vaccine introduction in India

**DOI:** 10.1101/2021.10.26.21265523

**Authors:** Syed F Quadri, Amanjot Kaur, Arindam Ray, Seema Singh Koshal, Mayank Shersiya, Pradeep Haldar, Sanjay Kapur, Mohammed Haseeb, Arup Deb Roy

## Abstract

**Background:** Studies have shown that while training has significant potential in improving the knowledge and skills of healthcare staff, the beneficial effect of training may decline with time. Studies have emphasized the role of assessment in understanding the relevance of the training structure and content and employing course correction as per the need. Besides, there is a lack of literature on the assessment of the level of knowledge retention among the participants. This study aims to conduct an assessment study to determine the level of knowledge retention.

**Methods:** The study was conducted among frontline health workers who received training on the rotavirus vaccine introduction. Assessments were conducted at a gap of one month and twelve months after the intervention. Simple percentages were used to compare the level of knowledge retention and McNemar’s chi-square test to determine P values.

**Results:** This is the first study conducted in India to assess the retention of knowledge 12 months after the new vaccine introduction training of health care professionals. The study comprised 41 participants who demonstrated an increase in the level of knowledge retention at the end of 1-month; however, a decline in the knowledge retention was seen at 12-months. For the issue of missed second dose among children who received the first dose, an increase in knowledge level and retention was observed.

**Conclusion:** The study results showed that a time-bound decline in knowledge retention occurs. The authors recommend regular monitoring, refresher training to supplement the primary training.

## Introduction

It is an undebatable fact that health care staff plays a key role in improving vaccination coverage among children. The knowledge and attitude of health care staff drive the frequency with which vaccines are delivered and accepted amongst the public (Anastasi et al., 2009)(LP et al., 2010). While it has been established that training of health care staff on immunization leads to an improvement in the knowledge of the health care personnel and has a positive impact on the vaccination uptake (AB et al., 2015; E et al., 2008; P & P, 2012; RB et al., 2015), it has also been seen that one-time training elevates the knowledge level of the attendees exponentially, but a substantial level of knowledge goes down by 6 to 12 months after training (Ameh et al., 2018; Bardosono et al., 2018; Draiko et al., 2019; Gass & Curry, 1983; NC et al., 2019; Nelissen et al., 2015; Offiah et al., 2019).

Available evidence has indicated that several assessment studies have been conducted among health care workers to evaluate the retention of knowledge on various topics, including helping babies breathe (Draiko et al., 2019), resuscitation (Govender et al., 2010), drug-resistant tuberculosis management(Wu et al., 2019), however, not too many assessment studies on the retention of knowledge on routine immunization have been conducted till date. A study from Nigeria assessed the impact of training intervention on immunization providers’ knowledge and practice of routine immunization three months and six months after the training intervention. The study results suggested a need for an ‘effective monitoring and supportive supervision system’ to ensure that the intended outcomes of the training are being met on-field (Brown et al., 2017). However, no such long-term assessment studies have been reported from India. Hence, there is a need to conduct assessment studies in India to evaluate the long-term knowledge retention level of health care staff after the training intervention.

The introduction of the rotavirus vaccine in India commenced in 2016, and through 4 phases, the introduction was completed across the country in 2019. During the introduction, seven trainings of trainers (ToTs), four regional trainings, and three state training were conducted for state immunization officers, cold chain managers, district immunization officers, and medical officers. A pool of 778 master trainers was created through the training through the Indian states and union territories. A standardized training module was developed and used for conducting the training across all the training sessions. During the training, the participants’ knowledge was assessed through a pre-test and post-test performed before and after administering the training module (Kaur et al., 2021).

While rotavirus vaccination has been successfully introduced across India, it is important to assess whether continuing training will play an important role in the health care staff’s knowledge base and skill level, thereby influencing behavior change and increased rotavirus vaccine coverage. In this article, the authors have conducted a telephonic assessment of the health care staff who were imparted training on rotavirus vaccination, at 6-months and 12-months following the training intervention. This study is the first study to assess the impact of a new vaccine (rotavirus vaccine) introduction training in frontline health workers in terms of retention of knowledge acquired after the training. The assessment results will help highlight the level of knowledge retention, the gaps in the existing training curriculums and delineate the need and frequency of refresher courses to sustain the knowledge gained at the time of administering the training curriculum.

## Methods

### Study participants

The study participants comprised the frontline healthcare workers (FLWs) working at the district level who were imparted training on rotavirus vaccine introduction. All the FLWs were working for the Government health facilities as part of the frontline immunization workforce.

Only those participants who were willing to enroll in the study gave verbal consent for the same and attended the assessment after one month of training were included in the study. All the participants were selected through simple random sampling from the district-wise list of participants. The participants were also asked if they agreed to participate in the assessment. Due to logistical reasons and the consent to partake in the study, 5 participants were included from each district.

Those participants who participated in the first round of the assessment but were not willing to participate in the second assessment were excluded from the study. The details of the training are described elsewhere.

### Study tools

The level of retention was assessed through a closed-ended questionnaire. The questionnaire was first developed in the English language and then translated into the regional language. Table 1 shows the survey tool used during the study. The questionnaire consisted of five questions covering all aspects of Rotavirus training, vaccination schedule, doses to be administered, location of vaccine vial monitor (VVM), and interchangeability of doses if the child misses a dose and comes back after one year. The interview was conducted telephonically.

**Table 1:** Survey tool

| **S. No** | **Questionnaire for District participants** | **Answer** |
| --- | --- | --- |
| **1** | What is the schedule of Rotavirus vaccine | 6, 10, 14 weeks |
| **2** | What is the dose of Rotavirus vaccine available in your state | 5 Drops |
| **3** | Where is the VVM on Rotavirus Vaccine vial | On the cap |
| **4** | If the child has taken 1st dose of the freeze-dried RVV from another state and attends an RI session in your state for 2nd dose, what should the health provider do? (Whether administer RVV available with him/her or not) | The child will complete the remaining doses with the type of vaccine available in your state. No child should be left unimmunized |
| **5** | If the child takes 1st dose at six weeks and again comes after 12 months for 2nd dose, will the health workers give the subsequent doses of the vaccine | Yes, health workers will give 2nd dose and ask the parents to come for 3rd dose after four weeks |

### Study design

The study is an interventional study with follow-ups conducted at six months and 12 months. The study was conducted in 14 districts of Punjab state in northern India. The assessment of the knowledge retention was made by the proportion of correct responses received from each respondent. The evaluation was conducted twice after training, once after a month’s gap, to check the participant’s knowledge before introducing the new vaccine. The second assessment was conducted after a gap of 12 months with the same participants to assess the level of retention. After the telephonic survey, the respondents were provided with the correct answers to reinforce or refresh their knowledge, as the need may be.

### Statistical analysis

The data were analyzed using the SPSS Version 20. Each correct answer scored 1 point, whereas the wrong and un-attempted answers were assigned a score ‘0’. Simple percentages were used to determine the decrease/increase in the level of knowledge gained after one month of training and at the follow-up of 12 months. McNemar’s test was conducted to get the P values.

### Ethics approval

The ethics approval for the study was obtained from the Institutional Review Board of MGM-ECRHS, MGM Medical College, Aurangabad, Maharashtra. The study was given ethical clearance to be conducted among selected study participants.

The assessment study is a Punjab government-initiated initiative. Verbal consent was taken from each participant before the commencement of the study. The data collected and recorded from all the participants was kept anonymous.

## Results

A total of 41 participants attended both the assessment interviews. Those participants who did not participate in the second assessment were not included in the analysis of the study. While most of the questions depicted a drop in the level of retention of knowledge acquired at the training session, the respondents demonstrated an upward trend in the level of knowledge for the question on the subsequent doses of vaccine. (Appendix 1) Figure 1 shows the comparative scores at one month and 12 months follow-up after the training.

**Figure 1:**
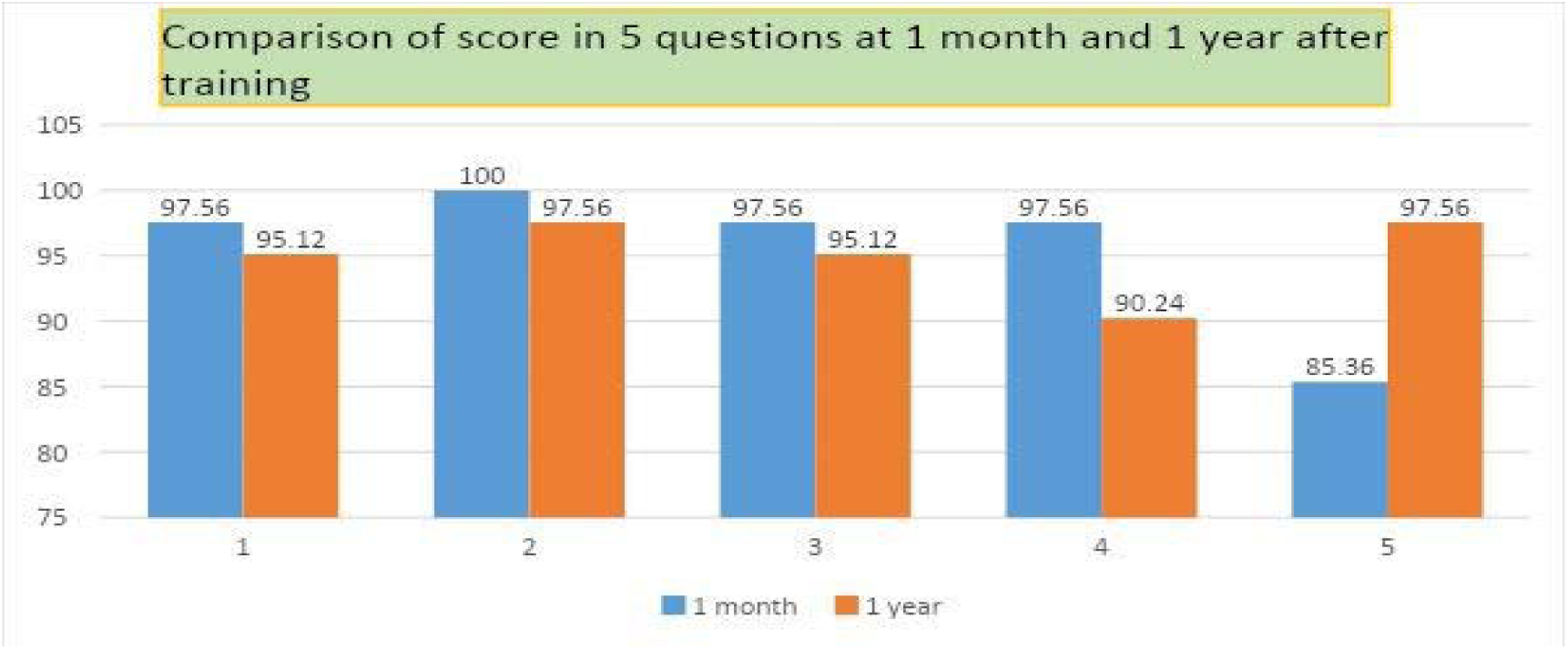
Comparative scores at one month and 12 months follow up

The results showed an overall reduction in knowledge retention. Table 2 depicts the participants’ performances and level of knowledge retention at one month and 12 months after the training. The staff showed a difference in the level of knowledge after a gap of 1 year in the absence of a refresher course.

**Table 2:** Comparative performance of the trained health care staff at 1-month and 12-months of the training intervention.

| **Question Number** | **At one month**<br>**N (%)** | **At twelve months**<br>**N (%)** | **P value (McNemar's chi square test) 1-tail** | **2-tail** |
| --- | --- | --- | --- | --- |
| **1** | 40 (97.6%) | 39 (95.1%) | 0.5 | 1 |
| **2** | 41 (100%) | 40 (97.6%) | 0.5 | 1 |
| **3** | 40 (97.6%) | 39 (95.1%) | 0.5 | 1 |
| **4** | 40 (97.6%) | 37 (90.2%) | 0.18555 | 0.3710 |
| **5** | 35 (85.4%) | 40 (97.6%) | 0.06529 | 0.1305 |

## Discussion

The current study demonstrates a deterioration of knowledge level as the time gap increases after the training was administered. In the present case, there was a time gap between the training and the initiation of the RVV vaccination program. In this period, no refresher training was provided to the staff after the initial training, and they started the vaccination program. Occasional monitoring and evaluation of the vaccination sites were undertaken by the state health officials and the partner organizations. While it was seen that most of the staff could carry on the vaccination sessions, a slight decrease was observed in the retention of acquired information gained at the primary training. The study results are in sync with previous studies conducted to assess the level of knowledge retention. A study conducted to determine the retention of knowledge and skill of birth attendants in newborn care and resuscitation demonstrated a significant reduction in the level of knowledge retention (21-77%) in clinical practice one year after the training was conducted. The study results also showed that after three months of training, substantial improvements were noticed in the proportion of knowledge gained in nutrition and parenting topics. However, a decline was detected at the end of three months compared to immediately after the training (Bardosono et al., 2018). Another study showed that following four weeks of intensive training of resident doctors, the optimum acquisition and retention of knowledge lasted for six months (GB et al., 2019). In this study, the maximum difference was noticed on the issue of vaccine interchangeability (7%). One reason that could be attributed to this knowledge loss may be that health care workers did not come across too many cases where people would have migrated between states using different vaccines. Hence, a lack of practical experience may have additionally affected the sustainability of knowledge gained and retained.

Interestingly, it was also seen that an increase in knowledge level (11%) occurred on the topic of missed dose and age up to which the second dose can be administered. The effect of on-site monitoring can explain this reverse trend, and the frequently asked questions (FAQs) booklet often referred to by the participants. Besides, the number of such cases may have been comparatively higher at the end of 12-months to have led to a clinical experience-based knowledge enhancement. Similar results were noticed in a study conducted to assess the knowledge retention on community maternal and newborn health among frontline health workers in rural Ethiopia at the 18-month follow-up (AG et al., 2014). Some other studies also demonstrated a sustained level of knowledge retention months after the training (Malau-Aduli et al., 2019; Sankar et al., 2013), followed by a gradual decline (Kaczorowski et al., 1998; WA et al., 2009). In a study among pre-service and in-service nurses, it was seen that the acquisition of knowledge and skills improved with training in both pre-service and in-service nurses. However, nurses with better exposure to patients due to being in service retained knowledge to a better level than pre-service nurses (Sankar et al., 2013). Another study also demonstrated an increase in overall performance levels with the increasing year of study. The study also emphasized the role of assessments in evaluating the relevance of the course structure and assisting in benchmarking knowledge retention and comparing results of the evaluation (Malau-Aduli et al., 2019). In terms of decline in knowledge, it was seen in a study conducted among residents that a deterioration in both neonatal skills and knowledge was observed, which did not show any improvement when the training was supplemented with the use of mannequins of booster videos on the training topics (Kaczorowski et al., 1998). In another study conducted among nurses on neonatal resuscitation programs, it was seen that training led to a substantial improvement and enhancement of knowledge and skills of neonatal resuscitation. However, after six months of training showed that there was a significant decrease in written and performance scores (WA et al., 2009). Our study has also demonstrated similar results, which are in sync with the results shown in earlier assessment studies.

The authors recommend that following the primary training, supplementary training should be conducted and ready reckoners in the form of FAQs, posters, and flipcharts may be redistributed digitally and in printed formats. Gobezayehu AG et al., in an earlier study conducted among the frontline health workers, suggested that refresher training, mentoring, and development of approaches to increase knowledge retention on issues where low performance is detected (AG et al., 2014). Regular session monitoring through inspection of the vaccination sites and assessment of trainers through interviews can also help assess the level of knowledge and skills gaps existing and bridge the gap with supplementary training. Hence, a capacity-building policy should reinforce a three-pronged approach to ensure knowledge retention and recall; primary training, monitoring, and repeat training (Figure 2).

**Figure 2:**
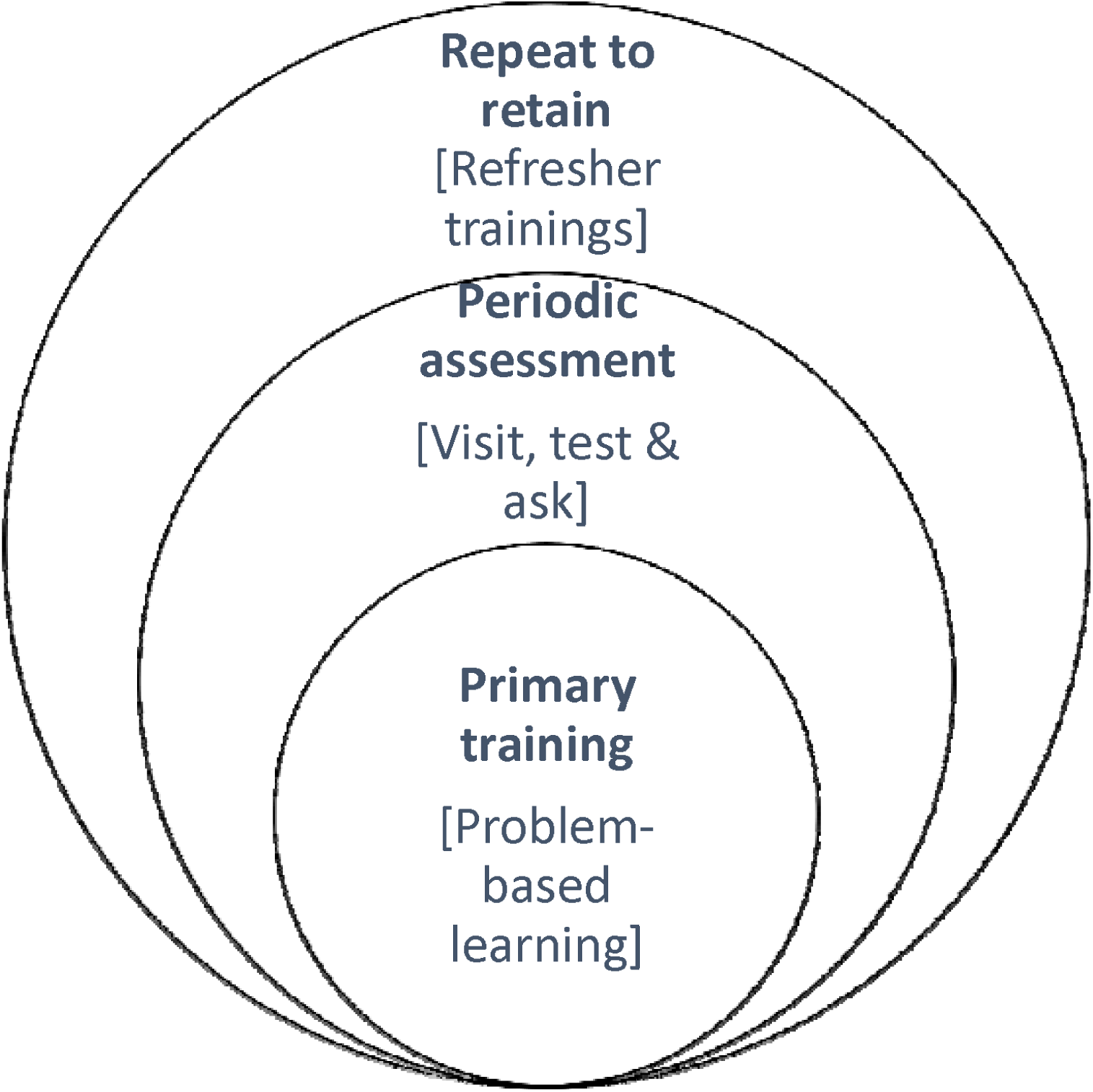
The three-pronged approach to training

## Study limitations

We conducted the assessment one month and 12 months after the training; hence there is no intermediate data on the level of knowledge retention of the participants. An additional evaluation at the end of 6 months of training would have helped track the trend in knowledge retention levels. Also, a lack of pre-test and post-test immediately before and after the training was not conducted, so we could not track the change in the knowledge level immediately back and after the training.

## Conclusion

Through earlier studies, it has been established that training interventions among health care professionals, with or without prior exposure to the topic under discussion, leads to an immediately high knowledge and skills acquisition following the training. However, there is a gradual decline in knowledge retention and deterioration of acquired skills with increasing time. Our study showed a significant increase in the knowledge level at the one-month assessment of the training, while a significant decline was seen at the 12-month assessment. The authors suggest that there should be some modification in the strategy adopted for capacity building and imparting the training. The authors recommend that problem-based learning, continuous monitoring, and refresher training should be made integral to the training structure.

## Data Availability

All data produced in the present work are contained in the manuscript

